# Real-World Changes in Movement-Evoked Pain and Gait in Adults With Knee Osteoarthritis

**DOI:** 10.64898/2026.05.11.26352918

**Authors:** Julien A. Mihy, Mayumi Wagatsuma, Spencer N. Miller, Elisa S. Arch, Katie A. Butera, Stephen M. Cain, Jocelyn F. Hafer

## Abstract

**Objective:** Adults with knee osteoarthritis often experience movement-evoked pain (MEP), and that pain has the potential to alter gait mechanics and influence disease progression. However, the associations between MEP and gait biomechanics have only been assessed in typical lab settings. Gait mechanics differ in the lab compared to in the real-world, thus it is unknown whether these associations between pain and gait translate to real-world settings. Therefore, this study aimed to measure concurrent changes in MEP and gait mechanics across three days of typical real-world activity.

**Design:** Seventeen participants with self-reported physician-diagnosed symptomatic knee osteoarthritis wore inertial measurement units on their more symptomatic limb’s thigh and shank, as well as on both feet for three days of typical activity. Participants were sent 5 automated text messages a day and were instructed to complete a short 3-5 minute walk and self-report their MEP via a Numeric Rating Scale (0-10) during each of the walks. A random coefficients model was used to determine how gait speed, stride length, and knee and ankle range of motion was related to changes in pain intensity.

**Results:** The average MEP experienced during the instructed walks was 1.4 ± 1.3 with individual participant average pain intensities ranging from 0 to 4.8. Greater MEP was associated with a 2.7° decrease in knee range of motion per unit increase in pain (95% CI [-4.8 -0.5], p = 0.02). Seven of the seventeen participants never reported a pain level of 0. Speed, stride length, and ankle range of motion did not differ by pain intensity.

**Conclusions:** Increases in MEP were associated with decreases in knee range of motion. A 2.7° decrease in knee range of motion in response to a 1-unit change in pain is meaningful as 5° is generally considered the threshold for a meaningful difference in joint angles. With a change in pain intensity of 2 being common with daily activity, individuals may be experiencing meaningful changes in knee joint angles regularly. With gait mechanics being associated with disease progression, these daily acute fluctuations in pain may be influencing disease progression rates.

## Introduction

Adults with knee osteoarthritis (OA) often experience movement-evoked pain, defined as pain with movement, and that pain influences gait mechanics. For example, larger joint moments and lesser walking speeds, stride lengths, and knee flexion angles have been tied to greater pain levels.^1–4^ To date however, associations between pain and gait biomechanics have only been assessed in typical lab settings.^5–11^ Gait mechanics differ in the lab compared to in the real-world,^12–14^ and so it is unknown whether there are similar associations between pain and gait in real-world settings. Real-world gait often includes slower walking speeds and shorter stride lengths than in-lab gait.^12–14^ Because speed and stride length are related to other gait mechanics, this may mean that joint ranges of motion and moments are also lower in real-world compared to in-lab gait.^15,16^ Based on cross-sectional studies,^1,3^ we might expect these smaller joint ranges of motion and moments in the real-world to be associated with less pain, but daily pain with walking and activity avoidance are common in those with knee OA.^17–21^ Therefore, our understanding of the association between pain and gait mechanics from in-lab studies may not translate to real-world gait.

Real-world gait is not only characterized by different gait mechanics than in-lab gait, but it also includes more variability in pain than what is generally captured during single in-lab visits.^12,13^ This variability in pain was captured by two studies measuring real-world momentary pain across multiple days which found meaningful within and between day fluctuations in pain.^19,21^ However, these studies did not capture concurrent pain and gait mechanics. Concurrent pain and gait have been captured by in-lab studies, but these studies generally only capture gait at a single pain level or after a single change in pain.^4–6^ Such study designs may not represent daily, real-world fluctuations in pain. Therefore, we do not know the variability of pain-related changes in gait during real-world walking. Measuring pain and gait mechanics concurrently throughout and between multiple days may provide insight into the association between varying pain and gait during typical activities of daily living. For clinicians, this insight may improve intervention design or recommendations for medication timing. For researchers, this insight may help improve study design and understanding of the variability in pain and gait response during or following activity.

Investigation of the relationship between pain and real-world gait is needed. Although complex to measure and interpret, real-world assessments provide insight into how participants perform when unobserved in their typical daily environment. Additionally, real-world assessment allows for the observation of within- and between-day fluctuations in gait and pain. Therefore, the purpose of this study was to evaluate how real-world gait differs in response to pain during three consecutive days of typical activity in adults with symptomatic knee OA. We hypothesized that greater pain levels would be associated with slower gait speed, shorter stride lengths, and less knee and ankle range of motion.

## Methods

### Participants

This project was a part of a larger study in which a power analysis was conducted where it was determined a sample size of 19 was needed to identify an interaction effect on the changes in gait mechanics and pain following bouts of activity between adults with and without knee OA.^22^ Therefore, nineteen participants (13 female, 62.6±4.3 years) with self-reported physician-diagnosed symptomatic knee osteoarthritis participated in this study. Only seventeen participants (11 female, 63.4 ± 3.5 years; Table 1) were included in the final analysis due to sensor failure.

**Table 1.** Demographic and KOOS (Knee Osteoarthritis and Injury Outcome Score) scores MEAN (SD). ADL – Activities of Daily Living, QoL – Quality of Life.

| **Knee Osteoarthritis (N=17)** |  |  |
| --- | --- | --- |
| **Sex** |  | 6M / 11F |
| **Age (yrs)** |  | 63.0 (3.9) |
| **BMI (kg/m<sup>2</sup>)** |  | 28.7 (4.5) |
| **KOOS** |  | 69.5 (14.6) |
|  | Pain | 74.5 (13.4) |
|  | Symptom | 71.2 (14.8) |
|  | ADL | 82.8 (16.0) |
|  | QoL | 59.6 (12.3) |

All participants met the American College of Rheumatology clinical criteria for knee OA by having the presence of occasional to frequent knee pain, minimal morning joint stiffness, and no palpable joint warmth. Participants were able to walk continuously for 30 minutes without assistive devices, had no knee joint replacements (arthroscopies >1 year prior were allowed) or current pain (besides the symptomatic knee), and had no cardiovascular, pulmonary, or neurological conditions that limited daily activity.

### Data Collection

Prior to real-world data collection, participants completed a campus visit consisting of an in-lab gait analysis followed by training on proper inertial measurement unit (IMU) placement and detailed instructions for their real-world gait assessments. In a previous study we aimed to assess concurrent gait and pain by collecting spontaneous real-world gait via IMUs and symptoms via text message. In that study, we found that participants often did not have gait data in the time windows when they answered pain texts. To ensure that we would have concurrent gait and pain measures in the current study, at the in-lab visit, we instructed participants to go for a ∼3-5 minute walk at their preferred speed when they received a text message, and then to answer the pain assessment questions in the text after their walk. Participants were also instructed to try to replicate a similar path each time (e.g. hallway at home or work).

Once participants demonstrated proper understanding of real-world data collection procedures, they were sent home with five IMUs (Opal v2, APDM/Clario, Philadelphia, PA), a charging dock, and an instruction manual for reference. Each day, participants placed IMUs on their symptomatic limb’s thigh and shank, as well as on both feet (Figure 1). The thigh and shank IMUs were placed on the lateral aspect of the midpoint of the respective segment using a strap. The foot IMU was placed in a pouch fixed to the shoelaces on the dorsum of the foot. A pair of custom slippers with an IMU pouch on the dorsum of the foot was given to participants to wear at home when shoes were not worn (Figure 1). IMUs collected continuous 3-axis accelerometer, gyroscope, and magnetometer data at 128 Hz in synchronized logging mode. IMU average battery life was ∼8 hours per day. Participants charged sensors each night and sensors were only collecting data while not charging.

**Figure 1.**
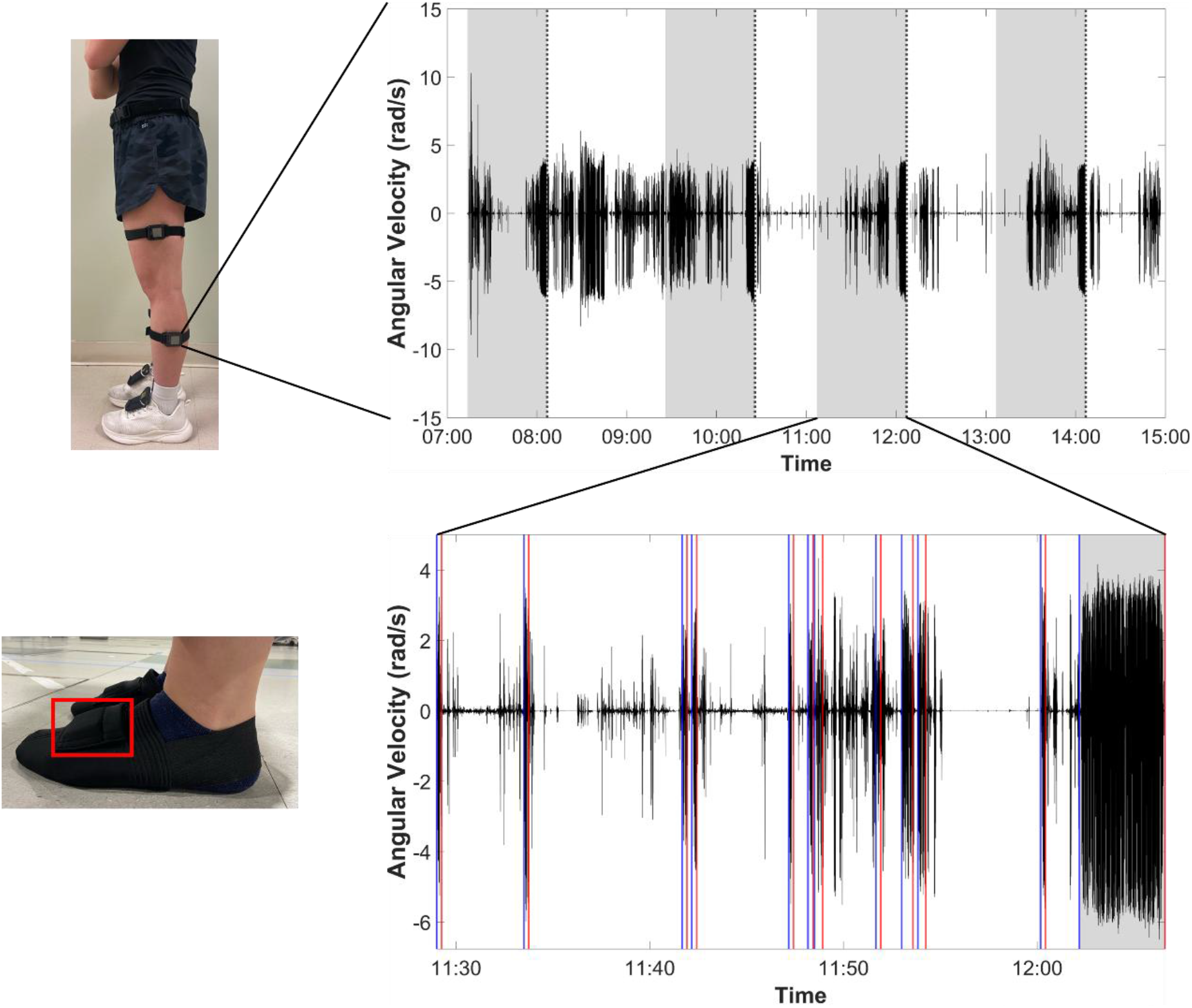
**Top left:** Example sensor placement. **Bottom left:** Slippers with sensor pouch worn at home if participants did not wear shoes. **Top right:** Shank IMU angular velocity data from one participant’s full day of wear. Vertical dotted lines represent when the participant completed the text surveys. Shaded regions show the hour range before a text survey response that was used to identify potential walking. **Bottom right:** Shank IMU angular velocity data from an identified hour range. Blue & red vertical lines represent the start (blue) and stop (red) times for each identified bout of walking. Shaded region shows the bout of walking used for analysis.

For each day of a three-day data collection, participants were instructed to don the IMUs when they woke up, go about their typical daily activities, and then dock the IMUs at night to charge. Data collection included two weekdays and one weekend day (Thursday-Saturday or Sunday-Tuesday). Participants received five automated text messages (Mosio, Seattle, WA) each day. The first text message was sent an hour after self-reported wake time and subsequent messages were sent every two hours. Each text message asked participants to follow a link to REDCap questionnaires that collected momentary movement-evoked pain levels. The questionnaires asked “What was your peak pain during the instructed walk?”. Pain was assessed on an 11-point numerical rating scale with 0 being no pain and 10 being the worst imaginable pain. The questionnaire link was available for one hour after the text message was sent. Responses to questionnaires were timestamped and these times were used to identify concurrent IMU data for gait analysis.

### Data Processing

Data processing began by selecting only the IMU data from the hour prior to each timestamped text message response (Figure 1). An hour window was used because, despite participant instructions, walking was not always identified immediately prior to the text. We then extracted bouts of walking within each IMU data segment based on repeated oscillations of the shank IMU. Raw shank IMU data were passed through a fast Fourier transform and 5-second windows with high-power density within frequencies representative of shank oscillations during walking (0.5-2.2 Hz) were considered walking. Consecutive 5-second windows with walking were combined.

All IMU data were oriented to a world reference frame, and each IMU’s functional mediolateral axis was determined.^23,24^ The world reference frame was created by aligning the vertical (Z) axis with gravity and making the mediolateral (X) axis and anteroposterior (Y) axis orthogonal in the horizontal plane. Alignment to the world reference frame was performed with a manufacturer-provided sensor fusion algorithm (APDM/Clario, Philadelphia, PA). The functional sensor-to-segment orientation was created by aligning the IMU mediolateral (X) axis with the functional axis of rotation during walking defined as the first principal component obtained when passing the IMU’s raw angular velocity data through a principal component analysis (pca function in MATLAB). Because IMUs may move throughout a day, separate functional mediolateral axis orientations were defined for each walking bout of interest. Data from the longest identified walking bout in the hour before each text were used for functional sensor-to-segment orientation.

Gait events were identified by passing the foot world-oriented vertical acceleration data through a one-dimensional continuous wavelet transform algorithm.^25^ The resulting wavelet peaks above a threshold (half the median magnitude of the peaks of the filtered signal) were defined as gait events. We used a ZUPT (Zero velocity UPdaTe) approach^26^ to identify periods of foot flat (i.e., zero foot velocity). We then integrated the foot world frame acceleration signal between periods of foot flat to calculate foot trajectories. Stride lengths, velocities, and directions were determined from foot trajectories between consecutive heel strikes. See Mihy et al. 2024 for more walking identification details.^27^

Segment excursions were calculated as the integral of the angular velocity about each segment’s functional mediolateral axis. Joint excursions were calculated for each stride as the difference between the segment excursions about that joint. The knee was calculated as thigh minus shank and the ankle as foot minus shank. Range of motion was calculated as the maximum joint excursion minus the minimum joint excursion for each stride.

The bouts used for analysis were the bouts of walking closest to each timestamped survey response that had at least 5 steady-state strides. Steady-state strides were defined as those with an inter-stride difference in stride length (m) and stride velocity (m/s) of less than 0.1.

#### Outcome variables

The outcome variables of interest were the peak pain values during the instructed walks, stride speed, stride length, and sagittal knee and ankle ROM. For each participant, outcome variables were averaged within each bout across the 3 days of data collection and each bout was included in the analysis.

#### Statistics

To determine how changes in pain relate to gait mechanics, JMP (v18.2.2) was used to perform a random coefficients model with up to 60 pairs (4 outcome variables x 5 time points x 3 days) using peak pain during the instructed walks as the predictor variable. The fixed effect was pain during the walks, random coefficients were the participants, and a residual repeated covariance structure was used (α<0.05).

## Results

A table of characteristics, pain, and gait kinematics for each bout used for analysis can be found at in Supplement Table 1.

### Bout characteristics

Between 2 and 12 timepoints were included per participant with an average of 8 per participant. Fewer timepoints than expected were captured for a few reasons. Surveys covered 10 hours of the day, but the IMU battery lasted roughly 8 hours and therefore the first or last survey timepoint often did not have concurrent IMU data. Timepoints were also missed because of either a lack of a survey response or lack of walking identified in the hour prior to a survey response. Bout duration was an average of 178 ± 215 sec and ranged from 20 to 1410 seconds.

The average time difference between the start of a bout and the corresponding text was 12 ± 15 min and ranged from 27 seconds to 59 minutes.

### Pain

Fifteen of seventeen participants (∼88%) experienced pain >0 at some point during the three days of typical activity (Figure 2 & Supplement Figure 1). The average pain experienced during the instructed walks was 1.4 ± 1.3 with individual participant average pain levels ranging from 0 to 4.8. Seven of the seventeen participants never reported a pain level of 0.

**Figure 2.**
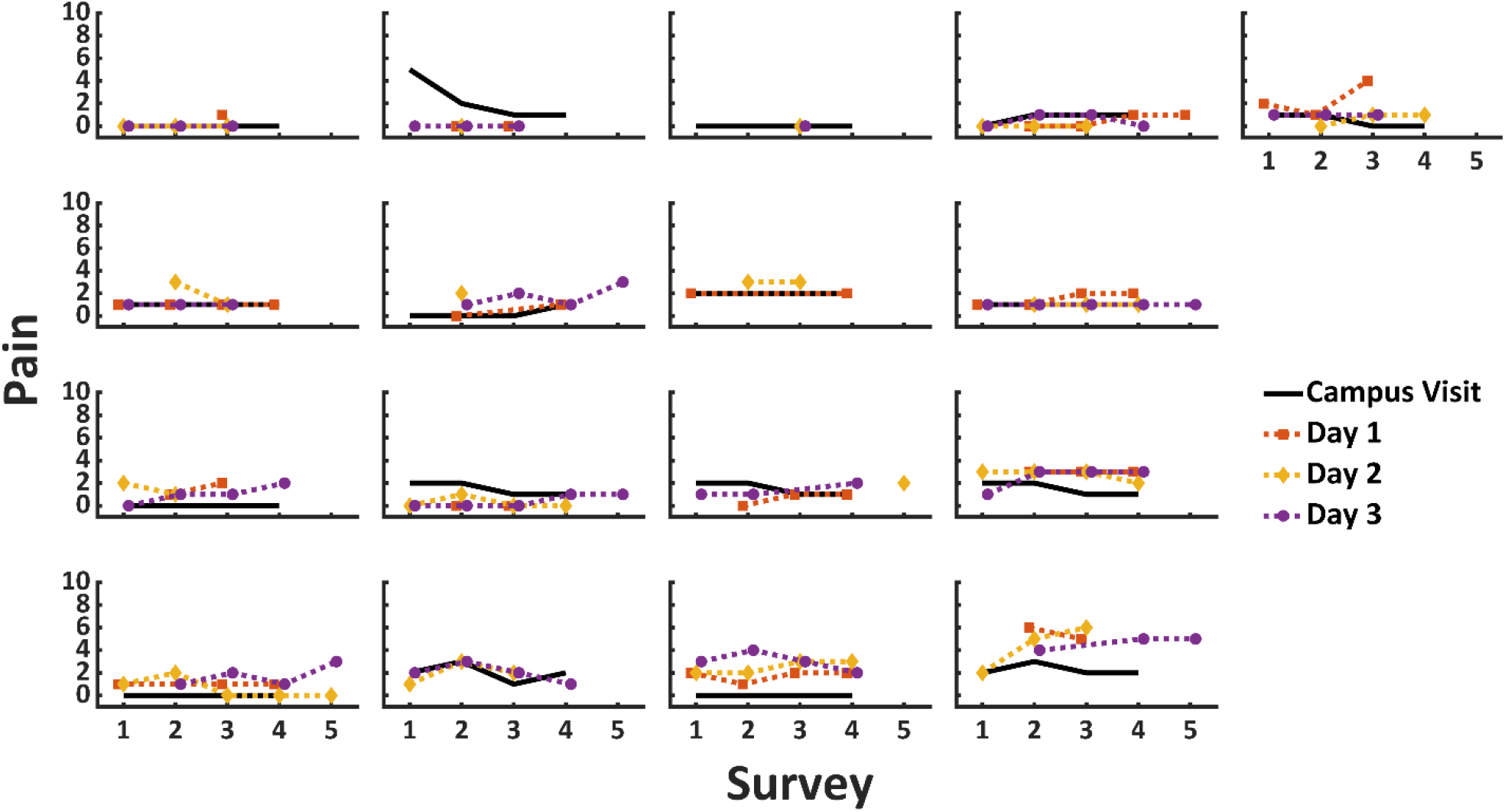
Peak pain during walking for each participant reported via text message surveys across 5 time points per day and 3 days. Days are represented by different line colors and marker shapes.

### Kinematics

Average stride speed across bouts were 1.1 ± 0.2 m/s and ranged from 0.5 to 2.8 m/s^2^. Average stride length within a bout was 1.2 ± 0.2 m and ranged from 0.7 to 2.1 meters. Average knee range of motion within a bout was 65.0 ± 11.8° and ranged from 91.8° to 37.9°. Average knee early stance range of motion within a bout was 11.6 ± 5.8° and ranged from 2.1° to 34.1°.

Average ankle range of motion within a bout was 35.0 ± 5.7° and ranged from 13.1° to 53.8°.

### Association between pain and kinematics

Greater pain was associated with less knee range of motion (95% CI [-4.8 -0.5], p = 0.02, Figure 3). The marginal model estimate identified a 2.7° decrease in knee range of motion per 1-unit increase in pain. The effect of pain across participants ranged from a decrease of 5.4° to an increase of 3.3° per 1-unit increase in pain. Speed, stride length, knee early stance range of motion, and ankle range of motion did not differ by pain level.

**Figure 3.**
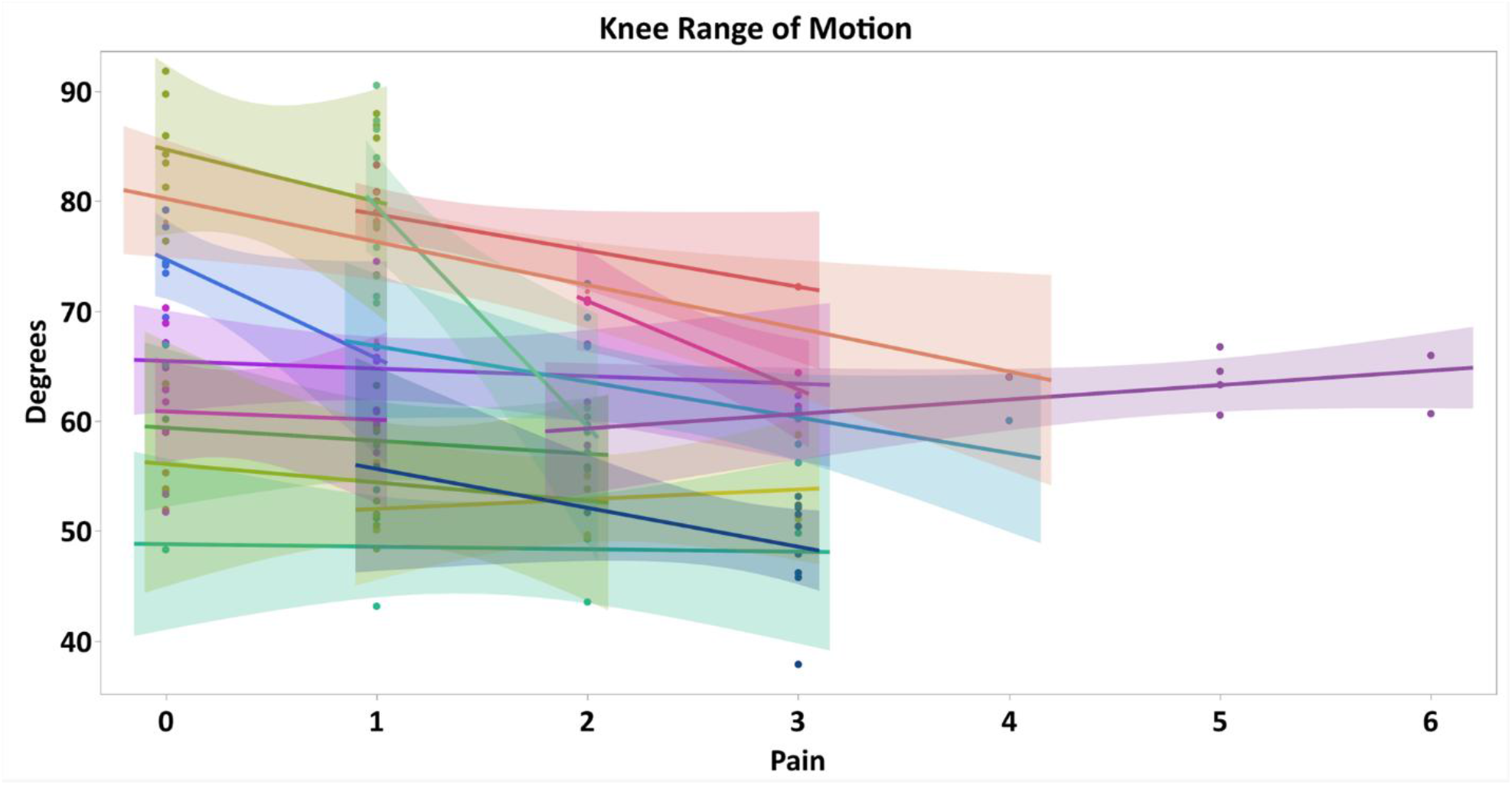
Relationship between pain and knee range of motion for all participants during three days of daily life. Each dot represents the average range of motion during a walk that had concurrent pain reported via text survey. Shaded regions show the confidence intervals for each line of best fit.

## Discussion

The aim of this study was to evaluate how real-world gait differs by pain level during typical activity in adults with symptomatic knee OA. Roughly 88% of participants experienced movement-evoked pain at least once during three days of data collection. Overall, movement-evoked pain ranged from 0-6 out of 10. Knee range of motion was the only variable to differ with pain, with a unit increase in pain being associated with a 2.7° decrease in range of motion. A 2.7° decrease in range of motion in response to a 1-unit change in pain is meaningful as 5° is generally considered the threshold for a clinically meaningful difference in joint angles.^28^ With a change in pain of 2 being likely with daily activity, individuals may be experiencing meaningful changes in joint angles regularly which may be influencing the rate of disease progression. This study highlights that individuals with knee OA alter their in knee range of motion in response to pain during unobserved real-world walking.

We observed fluctuations in pain across multiple days of typical activity and only knee range of motion differed with pain level. This aligns with Boyer and Hafer who found decreases in knee flexion in response to a pain-inducing protocol.^5^ They also found no changes in kinematics at the ankle. Based on in-lab studies measuring gait changes in response to pain relief, we also expected lower walking speed and stride length with greater pain levels,^4,29^ as these changes would decrease joint loads. However, the knee may be more sensitive to changes in pain than gait speed or the other measured variables.

This work was part of a larger study that also included a typical in-lab data collection with these same participants. During the in-lab data collection, participants completed ∼10-meter walks before and after multiple bouts of stairs aimed at inducing pain. In those data, we also only found that knee range of motion changed in response to bouts of activity despite no change in spatiotemporal variables (Supplement Table 1&2). This suggests that although in-lab gait may differ from real-world gait,^12–14^ individuals decrease their knee range of motion in response to pain regardless of setting.

A strength of real-world gait assessment and this study is the ability to measure fluctuations in pain across multiple days. In this study, 88% of the participants experienced pain during walking at some point over three days compared to 50% of the same cohort experiencing a change in pain during walking before and after a stair navigation protocol during an in-lab visit. Boyer and Hafer found ∼40% of participants experienced pain during an in lab treadmill protocol aimed at inducing pain.^5^ Not only did more participants experience pain in the real-world than in the lab, but 13/17 participants experienced greater pain in the real-world than in the lab (+1 to +3 greater maximum pain in the real-world, Supplement Table 1&2). It is often assumed that individuals with no change in pain in response to activity in lab studies have adopted a protective compensatory gait pattern. In-lab activity may not sufficiently capture other components influencing pain including day-to-day variations in function and symptoms. Therefore, the relationship between pain and gait may be misrepresented when only assessed during a single visit.

This study had some limitations and challenges. Two participants completed the study protocol but due to sensor failure, no IMU data were captured during the real-world portion. An additional limitation was IMU battery life. Because IMU batteries lasted an average of eight hours per day, not all text time points had concurrent gait mechanics. Therefore, our data skews towards collecting gait during the morning and early afternoon and may miss activity done by those that work and are more active in the evening. Our results may also be specific to our study design. When measuring real-world mechanics, we wanted to minimize our effect on participants’ typical activity so the only instruction given was to go on short walks when they received a text survey every couple of hours. This design led to some gait timepoints occurring immediately after an extended walk or other physical activity while other gait timepoints followed an extended period of inactivity. The variability in behavior before gait timepoints may have influenced pain levels and gait mechanics, but this does represent real-world behavior. Additionally, although participants were instructed to complete a 3-5 minute walk at each timepoint, the range of walking times was 20 seconds to 23.5 minutes. Although we analyzed the entire bout closest to each text message, bout duration influences stride speed and length and therefore may have influenced our findings.^14^

Measuring changes in gait in response to pain in the real-world is vital for determining how individuals with knee OA respond to the pain they experience while performing their typical activities of daily living. We found that higher pain levels were associated with less knee range of motion, but that spatiotemporal variables and ankle range of motion did not differ by pain level. As this project was part of a larger study, we were able to identify that these changes were similar to those found in the same cohort during an in-lab data collection that assessed walking before and after bouts of stairs. Therefore, participants responded to pain similarly between settings despite not being observed in the real-world and the pain being from daily activity as opposed to prescribed activity. This study demonstrates that individuals with knee OA consistently modify their knee range of motion as a compensatory method to decrease joint loads in response to increases in pain during activities of daily living in the real-world.

## Data Availability

All data produced in the present study are available upon reasonable request to the authors

## Notes

### Competing Interest Statement

The authors have declared no competing interest.

### Funding Statement

This study did not receive any funding

### Author Declarations

IRB of University of Delaware gave ethical approval for this work

