## Supplement Figure 1 for "Real-World Changes in Movement-Evoked Pain and Gait in Adults With Knee Osteoarthritis"

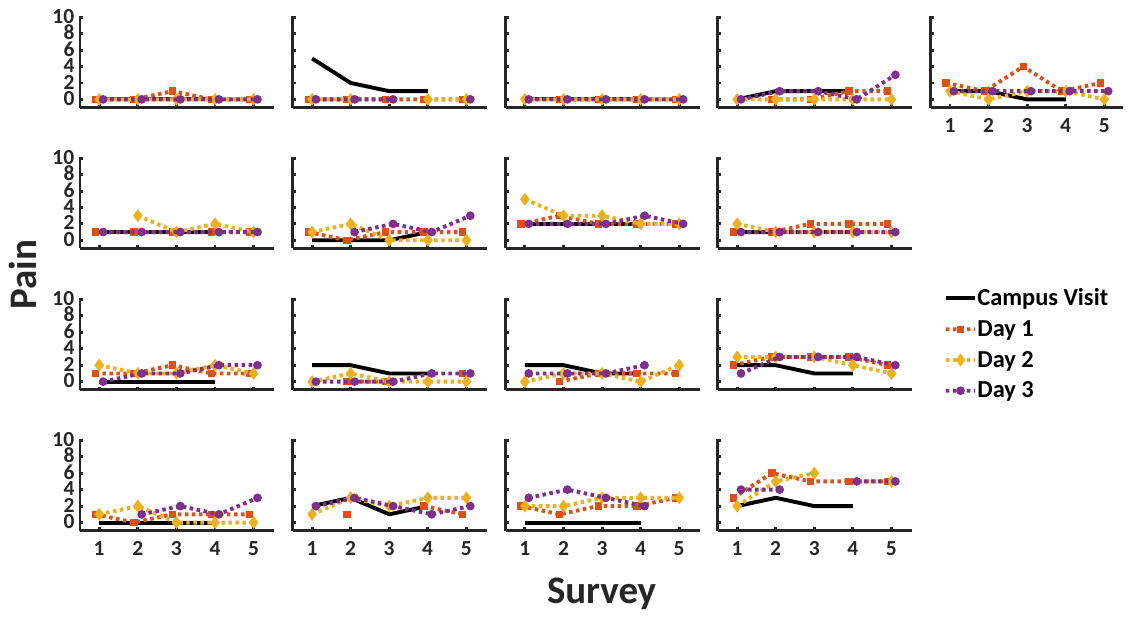

Peak pain during walking for each participant reported via text message surveys across 5 time points per day and 3 days. Days are represented by different line colors and marker shapes.
